# *ACE2* and *TMPRSS2* expression by clinical, HLA, immune, and microbial correlates across 34 human cancers and matched normal tissues: implications for SARS-COV-2 COVID-19

**DOI:** 10.1101/2020.04.29.20082867

**Authors:** Riyue Bao, Kyle Hernandez, Lei Huang, Jason J. Luke

## Abstract

**Background:** Pandemic COVID-19 by SARS-COV-2 infection is facilitated by the ACE2 receptor and protease TMPRSS2. Modestly sized case series have described clinical factors associated with COVID-19, while *ACE2* and *TMPRSS2* expression analyses have been described in some cell types. Cancer patients may have worse outcomes to COVID-19.

**Methods:** We performed an integrated study of *ACE2* and *TMPRSS2* gene expression across and within organ systems, by normal versus tumor, across several existing databases (The Cancer Genome Atlas, Census of Immune Single Cell Expression Atlas, The Human Cell Landscape, and more). We correlated gene expression with clinical factors (including but not limited to age, gender, race, BMI and smoking history), HLA genotype, immune gene expression patterns, cell subsets, and single-cell sequencing as well as commensal microbiome.

**Results:** Matched normal tissues generally display higher *ACE2* and *TMPRSS2* expression compared with cancer, with normal and tumor from digestive organs expressing the highest levels. No clinical factors were consistently identified to be significantly associated with gene expression levels though outlier organ systems were observed for some factors. Similarly, no HLA genotypes were consistently associated with gene expression levels. Strong correlations were observed between *ACE2* expression levels and multiple immune gene signatures including interferon-stimulated genes and the T cell-inflamed phenotype as well as inverse associations with angiogenesis and transforming growth factor-β signatures. *ACE2* positively correlated with macrophage subsets across tumor types. *TMPRSS2* was less associated with immune gene expression but was strongly associated with epithelial cell abundance. Single-cell sequencing analysis across nine independent studies demonstrated little to no *ACE2* or *TMPRSS2* expression in lymphocytes or macrophages. *ACE2* and *TMPRSS2* gene expression associated with commensal microbiota in matched normal tissues particularly from colorectal cancers, with distinct bacterial populations showing strong associations.

**Conclusions:** We performed a large-scale integration of *ACE2* and *TMPRSS2* gene expression across clinical, genetic, and microbiome domains. We identify novel associations with the microbiota and confirm host immunity associations with gene expression. We suggest caution in interpretation regarding genetic associations with *ACE2* expression suggested from smaller case series.

## Background

Severe acute respiratory syndrome (SARS) coronavirus 2 (SARS-CoV-2), which causes the disease COVID-19, was initially described near the end of 2019 [1, 2] and has caused a global pandemic. SARS-CoV-2 is a positive-sense single-strand RNA virus related to the SARS and the Middle East respiratory syndrome (MERS) coronaviruses that have caused previous global health emergencies [3]. COVID-19 is characterized predominately by fever, cough, and pneumonia, with some patients presenting with diarrhea and other symptoms [4, 5]. Mortality rates are described as approximately ten times higher than seasonal influenza in some clinical sub-groups [6].

Angiotensin-converting enzyme 2 (ACE2) has been identified as the receptor for the SARS-CoV family [7], and the SARS-CoV-2 spike protein binds ACE2 on host cells with greater affinity than previous SARS-CoV [8, 9]. Type II transmembrane serine protease TMPRSS2 is the primary human protease that mediates spike protein activation on infected cells, facilitating viral entry via receptor-mediated internalization [9, 10]. Multiple physiologic roles are known for ACE2 impacting systems such as cardiovascular, nephrology, and immune [11] but perhaps most notably related to SARS-CoV-2, pulmonary, where ACE2 has been described to limit severe acute lung injury [12]. Analyses of ACE2 protein expression by organ system have suggested high levels in epithelia of the lung and small intestine, consistent with presenting symptoms of patients with COVID-19 [13]. However, these studies have not integrated analysis of TMPRSS2 and integrated analyses may better inform which organ systems express both genes and may be at greatest infection risk.

Gene expression studies by bulk RNA sequencing and single-cell approaches have attempted to delineate expression patterns of normal airway tract and other tissues [14-16]. These studies have suggested high *ACE2* expression levels on the epithelia of oral and airway mucosa as well as small intestine. *ACE2* has additionally been suggested as an interferon-response gene suggesting a complicated interaction between viral infection and host antiviral response [15]. Further, a report has been advanced suggesting that lymphocytes may directly be infected by SARS-CoV-2 [17], a finding reported with MERS as well [18], however of unclear clinical significance.

Patients with cancer may be at particularly high risk for SARS-CoV-2 infection and deleterious outcomes to COVID-19 disease. In a single hospital study from Wuhan, China patients with cancer made up 1% of the overall prevalence of COVID-19 [19], substantially higher than the overall incidence of cancer in the Chinese population at 0.29% [20]. Outcomes to COVID-19 appeared to be worse in patients with cancer with increased intensive care unit admission, mechanical ventilation, and mortality, especially those who had recently received chemotherapy or surgery [19]. A subsequent literature-based international meta-analysis of COVID-19 incidence in patients with cancer has suggested a prevalence of approximately 2.0% globally [21]. Particularly there is concern that patients being treated with cancer immunotherapy drugs may be at even a greater risk given the possible overlapping immune-related toxicities for checkpoint blocking antibodies with the pneumonitis and diarrheal syndromes seen in COVID-19. Multiple societies, including the Society for Immunotherapy of Cancer, have issued guidance for cancer care during the pandemic as well as the use of immunomodulatory agents such as anti-IL6 [22].

Several clinical associations have arisen from a smaller series of patients infected with COVID-19. Some include risk factors for poor outcomes such as elevated body mass index (BMI) [23] and diabetes [24] as well as possible associations with race [25]. More broadly, germline genetics and host immune status may have a substantial impact on anti-viral host defense [26], similar to what is seen with cancer immunotherapy [27].

To better inform considerations surrounding SARS-CoV-2 and COVID-19 in patients with cancer and more broadly in the general population, we performed an integrated analysis of *ACE2* and *TMPRSS2* gene expression across clinical, genetic, and microbiome domains. We identify novel associations with the commensal microbiota and confirm host immunity associations with gene expression. We suggest caution against over-interpretation of clinical or genetic associations from smaller case series noting that these are not strongly associated with *ACE2* or *TMPRSS2* gene expression. We hope these data may better inform clinical considerations surrounding risk stratification and prevention approaches.

## Methods

### Datasets

Sample metadata tables were downloaded from The Cancer Genome Atlas (TCGA) (Genomic Data Commons (GDC) portal: https://portal.gdc.cancer.gov). Out of 11,093 aliquots total, 10,732 were selected to keep one unique aliquot per patient per sample type, as illustrated in **Supplementary Figure 1**. The final cohort consists of 9,657 primary tumors, 367 metastatic samples (all from skin cutaneous melanoma (SKCM)), and 708 normal tissues from 10,038 patients across 34 tumor types (33 primary and one metastatic) (**Supplementary Tables 1** and **2**). Out of 708 normal tissues, 14 tumor types have 15 or more normal samples available, hence were included for statistical comparisons when applicable. The standardized, upper-quartile normalized, batch-corrected, and platform-corrected RNAseq expression of 20,531 genes in RSEM (RNA-Seq by Expectation Maximization)-quantified read count estimates were downloaded from PanCancer Atlas consortium studies (https://gdc.cancer.gov/about-data/publications/pancanatlas) and log2-transformed for further analysis. FPKM (Fragments Per Kilobase of transcript per Million mapped reads) quantification of gene expression as well as whole-exome sequencing (WES) BAM files were downloaded from GDC [28]. Demographic and clinical information were retrieved from the TCGA Pan-Cancer Clinical Data Resource (TCGA-CDR) [29]. BMI and smoking history were retrieved from legacy clinical files. To the author’s knowledge, only two tumor types had diabetes status information (pancreatic adenocarcinoma (PAAD) and uterine corpus endometrial carcinoma (UCEC)), which was retrieved from clinical XML files on GDC. 82% of PAAD and 7% of UCEC patients have diabetes status recorded. Hence only PAAD was included in analysis. Commensal microbiota abundance and viral presence of TCGA samples were downloaded from published studies [30, 31]. Single-cell RNAseq (scRNAseq) gene expression in malignant cells, immune cells, and normal cells were retrieved from nine studies consisting of patients diagnosed with cancer and healthy donors. Links to data files, single-cell cohorts, and bioinformatics software are provided in **Supplementary Table 3**. Data generated in this study are accessible on GitHub repository https://github.com/riyuebao/ACE2_TMPRSS2_multicorrelates.

### *ACE2* and *TMPRSS2* gene expression correlation and percentile calculation

The gene expression of *ACE2* (Entrez Gene ID 59272) and *TMPRSS2* (Entrez Gene ID 7113) was retrieved from the RSEM-quantified RNAseq data and used for all analyses described in this study. Spearman’s correlation was calculated between the expression of the two genes in tumor (*n*=10,024) and normal (*n*=708) samples across all tumor types and within individual tumor types. The expression percentile was calculated separately within each of the four analysis sets (*ACE2* in normal, *ACE2* in tumor, *TMPRSS2* in normal, *TMPRSS2* in tumor) following two steps. First, the median expression of *ACE2* or *TMPRSS2* was calculated within individual tumor types. Next, tumor types were ranked by the median expression of each gene from higher to lower, and the position of each tumor type in the ranked list was scaled to 0 to 100, hereafter regarded as “expression percentile” per tumor type, with smaller values indicating top-ranked tumor types. The same process was repeated for each gene in tumor samples (34 tumor types) and normal tissues (14 tumor types).

### Analysis of clinical correlates

The expression of *ACE2* and *TMPRSS2* was compared between designated clinical groups, split by age (younger (<65 years) / older (≥65 years) in tumor or normal), gender (female / male in tumor or normal), race (African American (AA) / Asian / White in tumor or normal), menopause (not post / post in tumor or normal), BMI (level 1 (<25) / level 2 (25-30) / level 3 (30-35) / level 4 (>35) [32] in tumor or normal), smoking history (never / light / heavy [33] in tumor or normal), tumor stage (I / II / III / IV in tumor), tumor grade (G1G2 / G3G4 in tumor). For tumor grade, G1 and G2 were collapsed to indicate low-to mid-grade (G1G2), and G3 and G4 were collapsed to indicate high-grade (G3G4). Within each clinical factor, sub-groups of < 15 samples were excluded. For each clinical factor, comparisons were performed across all tumors and within individual tumor types. For all tumor types, first, data were fitted into a two-way ANOVA model with tumor type and clinical group as variables plus the interaction between the two. Second, if more than two clinical groups exist, Tukey’s honest significance test (HSD) was used with the fitted ANOVA model for pairwise comparisons while controlling for type I errors. Within each tumor type, Tukey’s HSD was used with one-way ANOVA models when more than two groups are present; otherwise Welch Two Sample *t*-test was used. The list of clinical groups and statistical results are provided in **Supplementary Tables 4, 5**, and **6**.

### Analysis of HLA correlates

HLA-A, HLA-B, and HLA-C genotypes were identified for 9,559 patients from TCGA across 34 tumor types using OptiType (v1.3.2) with WES BAM files. We performed two levels of analysis. In the allele level analysis, considering each patient carries two copies of HLA-A, B, or C alleles, both copies were counted towards the total number (19,118) of A, B, or C alleles in the entire cohort. In the patient-level analysis, only one HLA-A, B, or C allele was kept as the final label to assign to each patient, with the priority determined by the lexicographical ranking of all alleles present in the entire cohort. For each patient, between the two copies of HLA-A, B, or C alleles, if one was ranked before the other, then the first one was assigned to the patient. The calculation of HLA prevalence calculation was performed at the allele level. The comparison of *ACE2* and *TMPRSS2* gene expression between HLA genotypes was performed at the patient level using two-way ANOVA, given that gene expression was estimated per sample.

### Analysis of immune gene expression signatures

Five immune responsive and suppressive signatures (interferon-stimulated genes (ISG), T cell-inflamed (Tinfl), myeloid, angiogenesis (angio), and transforming growth factor-β (TGF-β)) (**Supplementary Table 7**) were correlated with *ACE2* and *TMPRSS2* gene expression in tumor and normal tissues. The expression level of a signature was computed as the average expression of all genes in this signature after centering and scaling. Spearman’s correlation was calculated between each signature and *ACE2* or *TMPRSS2*. The full correlation metrics are provided in **Supplementary Table 8**.

### Analysis of immune and stromal cell subset correlates

FPKM estimates of RNAseq gene expression was used for quantifying enrichment of 64 tumor and stroma cell types using xCell (v1.1.0). xCell converts gene expression into rank-based metrics within each sample, hence normalization and batch correction were not required. To make data comparable across samples, xCell was run once using all samples (*n*=10,732). Spearman’s correlation was computed between the enrichment score of each cell population and *ACE2* or *TMPRSS2* expression. The full correlation metrics are provided in **Supplementary Table 9**.

### Analysis of viral-associated tumors

HPV, EBV, and HBV were selected for this study as those are the three most prevalent cancer-associated viruses in the cohort. Other viruses detected were excluded. Samples were set to HPV positive/negative by cutoff 10, EBV positive/negative by cutoff 5, and HBV positive/negative by cutoff 5 given previously recommended thresholds [31]. STAD, ESCA, LAML, and OV were reset to “negative” for HPV, EBV, and HBV after a manual inspection, which revealed no strong clinical support for viral presence in those tumor types. Within each tumor type, *ACE2* and *TMPRSS2* gene expression was compared between viral positive and negative tumors using Welch Two Sample *t*-test. Across all tumor types, two-way ANOVA was used to compare gene expression between viral positive and negative tumors with tumor type and viral group as variables plus the interaction between the two.

### Analysis of microbial correlates

The abundance of 1,093 genus-level microbial taxa was quantified from tissue RNAseq data after rigorous QC, batch correction, and contamination filtering, and normalized to 1 million reads to make data comparable across samples [30]. Seven hundred six normal tissues and 9,801 tumor samples were included in the analysis where data were available. Taxa were filtered to keep bacteria in analysis; viruses and archaea were excluded. Nine hundred fifty taxa present in at least 20% of samples were kept for statistical testing. Within each tumor type, Spearman’s correlation was computed between each bacteria taxon and *ACE2* or *TMPRSS2* gene expression in tumor and normal tissues. For each test, at least 15 samples with taxon abundance ≥ one were required. 75 taxa passed FDR-adjusted *P*<0.05 and Spearman’s ρ > 0.5 or < −0.5 in at least one pairwise correlation (**Supplementary Table 10**).

### LASSO regression modeling and variable importance

LASSO regression models of *ACE2* or *TMPRSS2* gene expression were built in tumor (*n*=10,024) and normal (*n*=708) samples separately using 90 features consisting of tissue type as well as clinical (age, gender, race), immune signatures (ISG, T cell-inflamed, myeloid, angiogenesis, TGF-β), immune cell subsets (macrophage M1, macrophage M2, CD8 T cell, CD4 T cell), stroma cell subset (epithelial cell), and 75 microbiota features from microbiota correlation analysis (**Supplementary Figure 2**). Macrophage M1/M2 and T cells were included in the model based on emerging evidence suggesting an important role of macrophage and proinflammatory phenotype [34] in COVID-19 disease. Thirty-four tumor types were collapsed into 15 tissue types based on categorizations from The Human Protein Atlas to reduce complexity. Categorical variables were converted to dummy variables using R function *dummyVars* with parameter fullRank set to TRUE. Data were preprocessed to remove features that have near-zero variance, high correlation (Spearman’s ρ > 0.75), or high collinearity. Each feature was scaled and centered. Given the purpose of this analysis was to evaluate the relative importance of features rather than training and validating a predictive model, we did not split samples into training and test sets. Instead, we used all samples with 10-fold cross-validation. Variable importance was reported as raw values and as scaled values to 0-100 (**Supplementary Table 11**). R package caret (v6.0-84) was used for analysis.

### Statistical analysis

In all analyses, a minimal sample size of 15 per group was required for statistical testing. For group-wise comparisons, two-way ANOVA was used with tumor type and a group of interest as variables plus the interaction. When more than two groups are present, Tukey’s HSD test was used for pairwise comparisons. Within each tumor type, two-sided Welch Two Sample *t*-test was used to compare log2-transformed gene expression between groups; for paired samples, two-sided paired *t*-test was used. Spearman’s correlation was used to determine the relationship between two continuous variables. For multiple comparisons, p-values were corrected using Benjamini & Hochberg (BH)-FDR method. LASSO regression models were used to evaluate variable importance with 10-fold cross-validation. Those analyses were performed using R (v3.6.1) and Bioconductor (release 3.10). P-values less than 0.05 were considered statistically significant.

## Results

### *ACE2* and *TMPRSS2* are highly expressed in digestive organs and tumors, however, lower in tumor compared to matched normal

Given the association between COVID-19 disease and various organ-specific symptoms, we investigated the distribution of *ACE2* and *TMPRSS2* expression in tumor and normal tissues across 34 tumor types consisting of 15 tissue types (**Supplementary Tables 1, 2**, and **3**). None of the acute myeloid leukemia (LAML) samples express *TMPRSS2*; hence 33 tumor types were used for the analysis of *TMPRSS2*.

When ranked by expression percentile in each tumor type, cholangiocarcinoma (CHOL), colon adenocarcinoma (COAD), pancreatic adenocarcinoma (PAAD), rectum adenocarcinoma (READ), and stomach adenocarcinoma (STAD) are among the top 25% percentile for both genes, all of which are digestive organs (**Figure 1A**). We acknowledge that *TMPRSS2* is highly expressed in prostate adenocarcinoma (PRAD) likely due to known *TMPRSS2*:*ERG* gene fusion overexpression [35]. *ACE2* and *TMPRSS2* expression showed weak positive correlation in normal (Spearman’s ρ = 0.28, FDR-adjusted *P*<0.0001) and in tumor (ρ = 0.30, FDR-adjusted *P*<0.0001) (**Figure 1B**). Correlation within each tumor type showed a consistent pattern and can be further explored in external data files available on GitHub (https://github.com/riyuebao/ACE2_TMPRSS2_multicorrelates).

**Figure 1.**
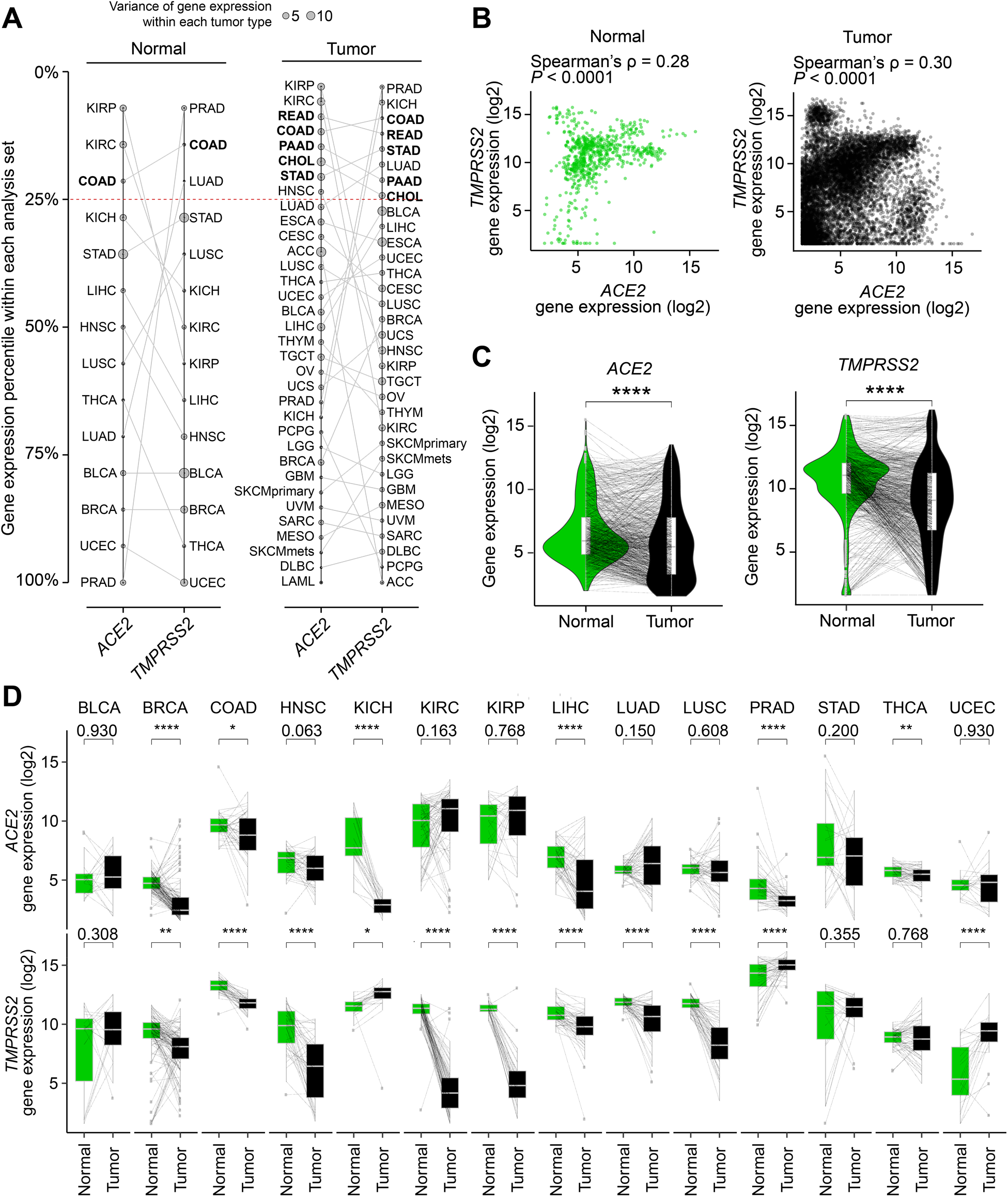
Distribution of *ACE2* and *TMPRSS2* gene expression in tumor and normal tissues across 34 tumor types. (**A**) Tumor types ranked by expression percentile in each gene. Five tumor types that show > 25% expression percentile within each analysis set in both genes are bolded. Four analysis sets are shown: (left panel) *ACE2* in normal (*n*=14 tumor types), *TMPRSS2* in normal (*n*=14 tumor types); (right panel) *ACE2* in tumor (*n*=34 tumor types), *TMPRSS2* in tumor (*n*=33 tumor types; LAML not shown due to lack of *TMPRSS2* expression in this tumor type). (**B**) Correlation between *ACE2* and *TMPRSS2* gene expression in normal (*n*=708 samples) and tumor (*n*=10,024 samples). (**C** and **D**) *ACE2* and *TMPRSS2* gene expression are higher in normal relative to tumor samples in (**C**) all tumor types pooled and in (**D**) individual tumor types. Line connects tumor and matched normal samples from the same patient (*n*=692 patients). Spearman’s correlation was used in **B**. Two-sided paired *t*-test was used in **C** and **D**. P-values shown are after FDR correction for multiple comparisons. \*\*\*\**P*<0.0001, \*\*\**P*<0.001, \*\**P*<0.01, \**P*<0.05. For comparisons that do not reach significance level of 0.05, exact p-values are shown. A full description of each cancer ID is provided in **Supplementary Table 1**.

In 14 tumor types where 15 or more matched normal tissues are available (**Supplementary Table 1**), we compared gene expression between tumor and matched normal from the same patients. The expression of *ACE2* and *TMPRSS2* was significantly higher in normal tissues relative to tumors (**Figure 1C**). Within individual tumor types, this pattern was significant for *ACE2* in breast cancer (BRCA), colon adenocarcinoma (COAD), chromophobe kidney cancer (KICH), liver hepatocellular carcinoma (LIHC), prostate adenocarcinoma (PRAD), thyroid cancer (THCA), and for *TMPRSS2* in 8 tumor types (FDR-adjusted *P*<0.05 and higher in normal). Three tumor types showed elevated levels for both genes in normal tissues compared to tumors (BRCA, COAD, and LIHC) (**Figure 1D**).

### Clinical factors and HLA genotypes do not strongly associate with *ACE2* and *TMPRSS2* gene expression in tumor or normal tissues

Several clinical observations on COVID-19 indicated clinical factors such as BMI might be associated with severity [5, 23]. We sought to investigate the association between clinical variables (age, gender, race, tumor stage, tumor grade, menopause, BMI, smoking history) and *ACE2* or *TMPRSS2* gene expression in tumor or normal tissues from 10,038 patients. For each clinical factor, data were fitted into a two-way ANOVA model with tumor type and clinical factor as variables plus the interaction between the two. Overall we did not observe significant differences in expression for either gene when comparing designated clinical groups within tumor or normal tissues (**Figure 2A** to **2F**) (**Supplementary Tables 4** and **5**). Outliers exist though are of unclear clinical relevance at this time (**Supplementary Table 6**). The results suggested those clinical variables are not strongly associated with *ACE2* or *TMPRSS2* expression. In addition, we investigated the association between the presence of diabetes and gene expression in pancreatic adenocarcinoma (PAAD), where data were available. Similarly, no significant differences in gene expression were detected in tumor or normal tissues from patients with or without diabetes.

**Figure 2.**
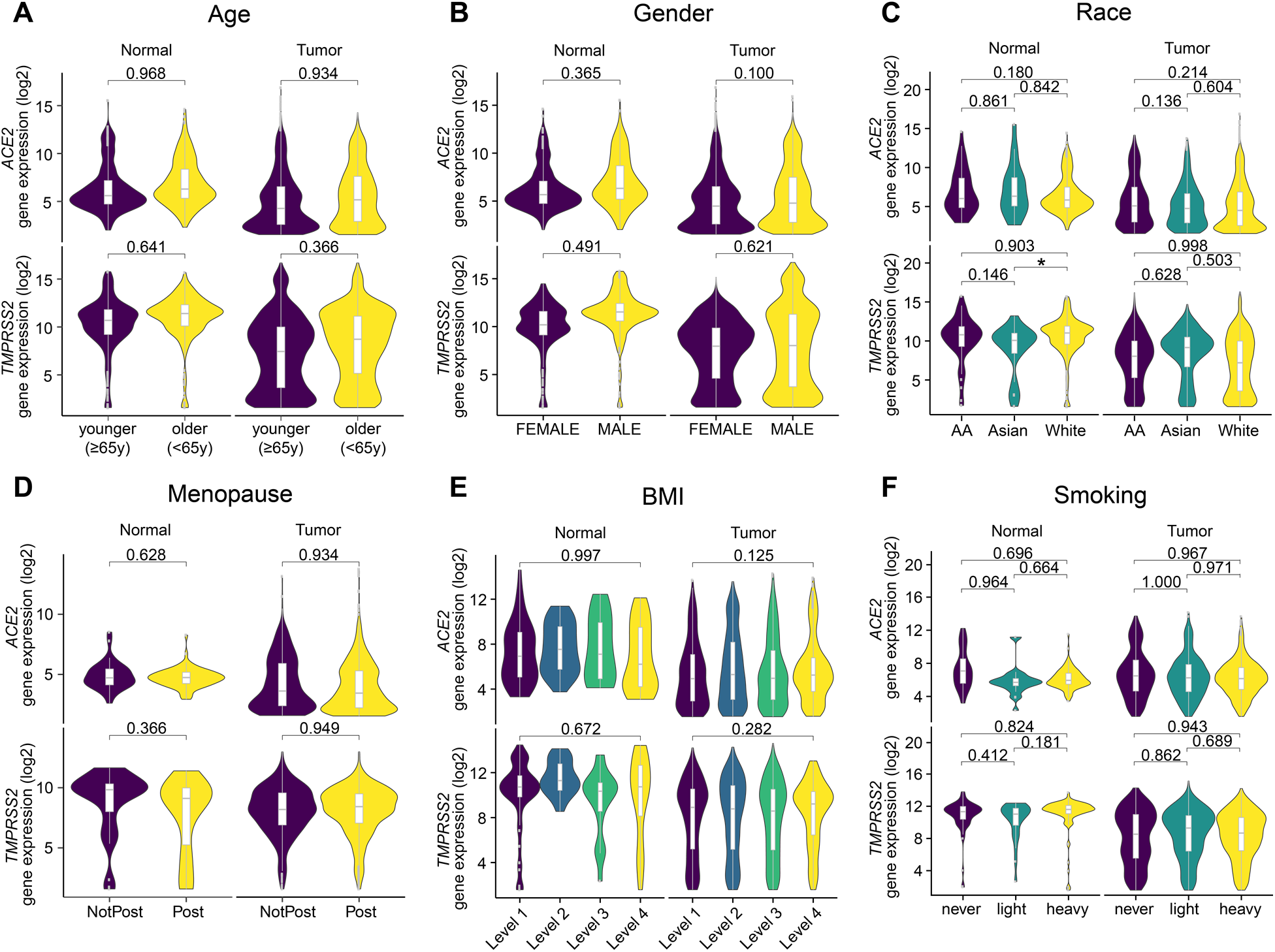
Clinical correlates of *ACE2* and *TMPRSS2* expression do not show consistent patterns across individual tumor types. (**A** to **F**) Gene expression by age (<65y, ≥ 65y), gender (female, male), race (African American, Asian, White), menopause (non-post, post), BMI (levels 1 to 4), smoking history (never, light, heavy), and by sample type (tumor, normal) when applicable. The criteria for each group definition are described in **Methods**. The number of samples in each group is provided in **Supplementary Tables 4** and **5**. In **E**, only p-values between levels 1 and 4 are shown; the rest is provided in **Supplementary Table 5**. Two-way ANOVA was used in **A** to **F**, with tumor type and clinical group as the variables plus interaction between the two. For clinical factors that have more than two groups (**C, E, F**), Tukey’s honest significance test (HSD) was used with the fitted ANOVA model for pairwise comparisons while controlling for Type I errors. Two-way ANOVA p-values after BH-FDR correction are shown in **A, B, D**, and Tukey’s HSD p-values are shown in **C, E, F**. \**P*<0.05. For comparisons that do not reach significance level of 0.05, exact p-values are shown.

Two HLA genotypes (B*46:01, B*54:01) have been reported to be associated with severe clinical outcomes by other groups [36]. We investigated the prevalence of the two alleles and identified low prevalence across all tumor types (0.6% and 0.2%, respectively, out of 19,118 HLA-B alleles from 9,559 patients). When looking into individual tumor types, both alleles were found to be significantly enriched in liver hepatocellular carcinoma (LIHC; FDR-adjusted *P*<0.0001), which was likely due to the enrichment of Asian populations in this cohort (43%). Comparison of *ACE2* or *TMPRSS2* gene expression among 36 HLA-A, 44 HLA-B, and 25 HLA-C genotypes with 15 samples or more per genotype showed no significant differences after adjusting for tumor-type specific gene expression, and can be further explored in external data files available on GitHub.

### *ACE2* correlates with distinct immune gene expression signatures and cell subsets while *TMPRSS2* correlates with epithelial cell populations in tumor and normal tissues

To understand potential associations of *ACE2* and *TMPRSS2* in tissues relevant to patients being treated with cancer immunotherapy, we investigated the correlation between *ACE2* or *TMPRSS2* and immune gene expression signatures known to be relevant in immuno-oncology (**Supplementary Tables 7** and **8**). *ACE2* was positively correlated with ISG signature in tumor samples from 24/34 tumor types (71%, 14 reached FDR-adjusted *P*<0.05) and normal tissues from 10/14 tumor types (71%, 4 reached FDR-adjusted *P*<0.05), and negatively correlated with angiogenesis and TGF-β signatures (**Figure 3A**). *TMPRSS2* showed a mixed pattern of correlations with those immune responsive or suppressive signatures (**Figure 3B**).

**Figure 3.**
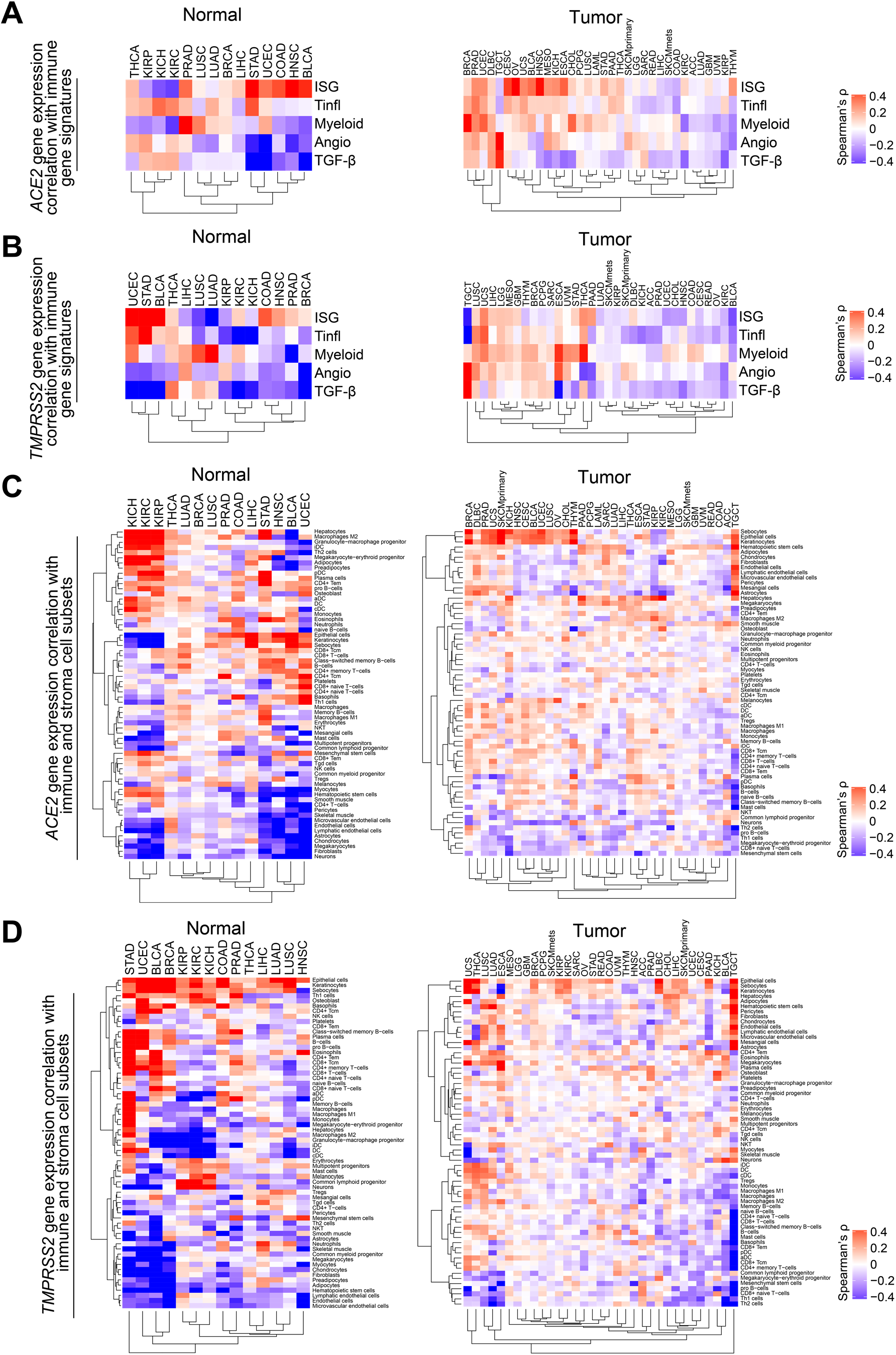
*ACE2* and *TMPRSS2* expression are correlated with distinct immune signatures or cell populations. (**A** and **B**) Correlation of five immune gene expression signatures, Interferon stimulated genes (ISG), T cell-inflamed (Tinfl), myeloid, angiogenesis (angio), and transforming growth factor-β (TGF-β) with (**A**) *ACE2* and (**B**) *TMPRSS2* gene expression. (**C** and **D**) Correlation of 64 immune, stroma, and other cell subsets with (**C**) *ACE2* and (**D**) *TMPRSS2* gene expression. *n*=14 tumor types shown for both genes in normal tissues. *n*=34 tumor types shown for *ACE2* in tumor, and *n*=33 tumor types shown for *TMPRSS2* in tumor. The full correlation statistics are provided as **Supplementary Tables 7** and **8**. Spearman’s correlation was used in **A** to **D**.

We did not find a consistent pattern of *ACE2* with cell subsets across all tumor types with the exception of a high positive correlation with macrophage M2 in normal tissues from kidney cancers (KIRC, KICH, KIRP) (Spearman’s ρ = 0.84, 0.82, 0.79, FDR-adjusted *P*<0.0001) and stomach adenocarcinoma (STAD) (ρ = 0.67, FDR-adjusted *P*<0.001) (**Figure 3C**) (**Supplementary Table 9**). *TMPRSS2* was positively correlated with epithelial cell abundance in tumor samples from 29/33 tumor types (88%, 17 reached FDR-adjusted *P*<0.05) and normal tissues from 14/14 tumor types (100%, 9 reached FDR-adjusted *P*<0.05) (**Figure 3D**).

With the observation of correlation with specific immune signatures, we sought to investigate whether *ACE2* or *TMPRSS2* were expressed directly by immune cells or other specific cell types in nine independent single-cell RNAseq studies. This includes tumor and immune cells from cancer patients diagnosed with glioblastoma or melanoma (Single Cell Portal, Broad Institute), or head and neck cancer [37]. In the cancer patient cohorts, less than 1% of the malignant cells express *ACE2* and/or *TMPRSS2*, while few to none of the immune cells express either gene. An exploration of three studies focusing on immune cells from healthy donors (13,316 PBMCs (Immune Cell Atlas), 39,563 ileum lamina propria immunocytes (Immune Cell Atlas), 594,857 immune cells (Census of Immune Cells, EBI)) further confirmed the lack of *ACE2* and *TMPRSS2* expression in immune cell populations. Realizing that other studies had reported the genes as highly expressed in lung, heart, brain, and colon, we investigated published large-scale profiling of 702,968 single cells from non-cancer patients or healthy donors (Human Cell Landscape) [38]. We found *TMPRSS2* was expressed in stomach, colon, kidney, prostate, intestine, jejunum, pancreatic, esophagus, and bladder tissues, while *ACE2* was only expressed in jejunum and fetal intestine. Lastly, we examined a public scRNAseq cohort from patients with COVID-19 infection (https://doi.org/10.5281/zenodo.3747336) and confirmed that *ACE2* was not expressed in immune cells, and *TMPRSS2* was present in six out of 140,956 cells total by one read count. Therefore, we concluded that *ACE2* or *TMPRSS2* are not expressed in immune cell populations, at least in the cohorts investigated. The expression of both genes in bulk RNAseq data was likely to be derived from non-immune cells, such as epithelial cells in the tissues.

### *ACE2* and *TMPRSS2* gene expression associates with microbiota in normal tissues particularly from colon and stomach adenocarcinoma

Given associations between strong anti-tumor immune responses due to the presence of tumor-related virus and particular commensal microbiota, we sought to investigate associations between *ACE2* and *TMPRSS2* gene expression with the presence of virus or tissue microbiota. Across known viral positive and negative tumor types, we found an inconsistent pattern relative to viral associated *versus* non-viral associated tumors (HPV, EBV, HBV). We then correlated 1,093 commensal microbiota identified in tumor and normal tissue RNAseq data, as published in [30] with gene expression. We identified 75 taxa that showed significant and strong positive or negative correlation (Spearman’s ρ > 0.5 or < −0.5) with either gene in at least one pairwise correlation (FDR-adjusted *P*<0.05) (**Figure 4A** and **4B**) (**Supplementary Table 10**). Colon adenocarcinoma (COAD) and stomach adenocarcinoma (STAD) were the two tumor types demonstrating the strongest and most prevalent positive correlation of *ACE2* and *TMPRSS2* gene expression with abundance of specific bacteria taxa, respectively. Kidney cancers, including renal papillary cell carcinoma (KIRP), chromophobe (KICH), and renal clear cell carcinoma (KIRC), also showed positive correlations between *ACE2* and microbiota if not as prevalent as colon or stomach cancers. If ranked by Spearman’s correlation coefficient, Chlamydia was the top microbiota positively correlated with *ACE2* in colon adenocarcinoma (ρ = 0.81, FDR-adjusted *P* < 0.0001) (**Supplementary Table 10**). For both genes, those patterns were less prominent in tumor samples, which could be due to high heterogeneity in tumors. Overall we observed approximately a 2.6:1 ratio of commensal microbiota for gram-negative to positive groups in the 75 taxa (50 negatives, 19 positives, others undetermined) (**Supplementary Table 10**).

**Figure 4.**
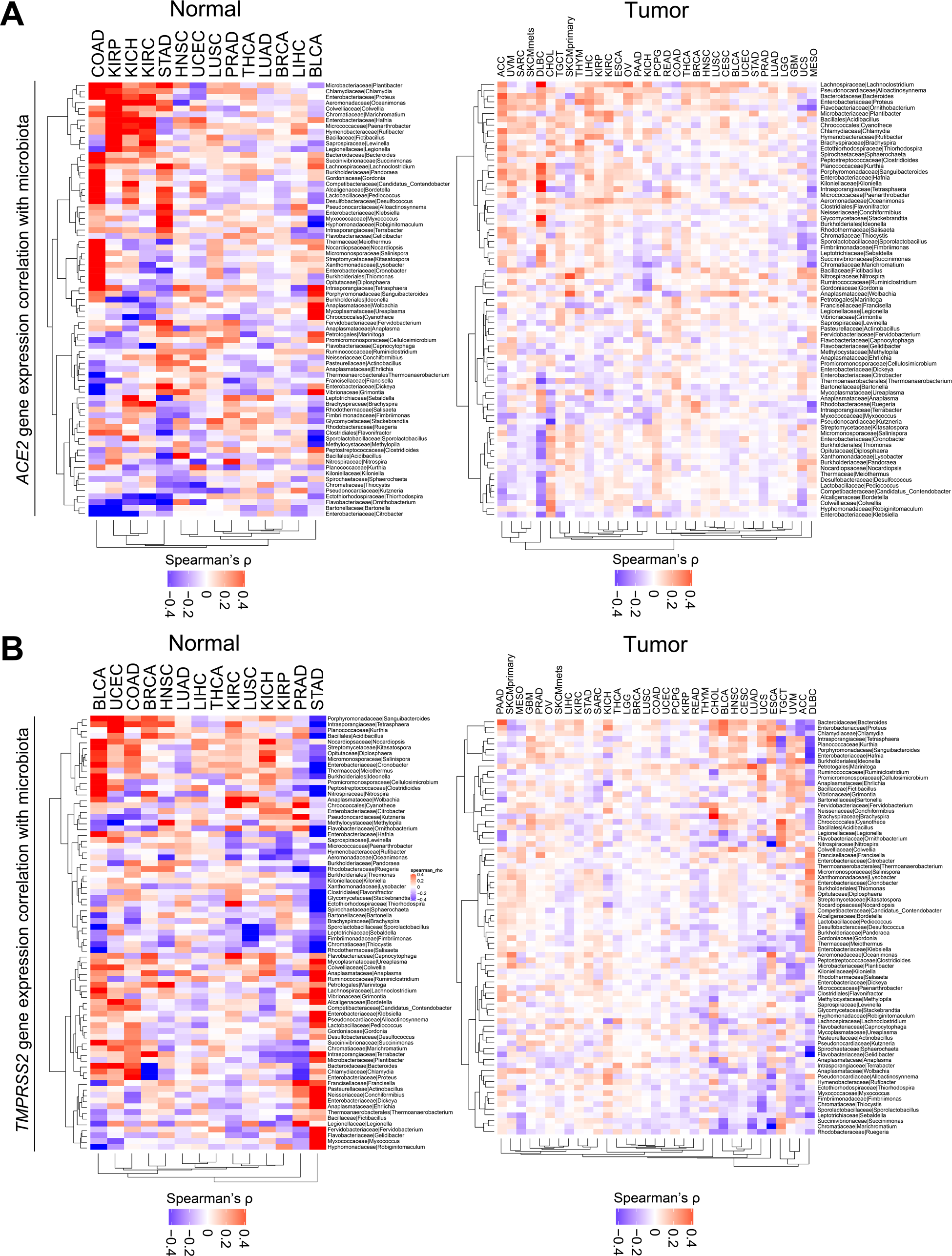
*ACE2* and *TMPRSS2* expression are correlated with distinct microbiota communities. (**A** and **B**) Correlation of 75 bacteria taxa with (**A**) *ACE2* and (**B**) *TMPRSS2* gene expression. Within each plot, the left panel shows normal tissues, and the right panel shows tumor samples. Seventy-five taxa were selected by correlation coefficient ρ > 0.5 or < −0.5 and FDR-adjusted *P*<0.05 in at least one pairwise correlation (**Supplementary Table 10**). For both genes, *n*=14 and *n*=33 tumor types are shown in normal and tumor samples, respectively. LAML does not have data available for microbiota abundance, hence excluded from analysis for both genes. Spearman’s correlation was used in **A** and **B**.

### Integration of multi-dimensional correlates revealed specific contributors shaping *ACE2* and *TMPRSS2* expression in tumor and normal tissues

To integrate all correlates and evaluate their relative importance in determining the gene expression of *ACE2* and *TMPRSS2*, we built LASSO regression models in tumor and normal tissues separately utilizing features from the clinical, immune, and microbial domains (**Supplementary Figure 2**). Clinical features included were age, gender, and race, while menopause, BMI, and smoking history were excluded because >50% of the samples were missing information. HLA genotype was not included because of many categories and/or levels, which may lead to overfitting. Immune gene expression signatures included ISG, T cell-inflamed, myeloid, angiogenesis, and TGF-β. Immune cell type features included macrophage M1/M2, CD8, and CD4 T cells, and non-immune cell type features included epithelial cells. Microbe features included the 75 bacteria taxa from the analysis above. In addition, we collapsed 34 tumor types into 15 tissue types and included these in the model to account for tissue-specific gene expression variations.

We calculated the importance of each feature in the models with 10-fold cross-validation (**Supplementary Table 11**). After quality control and filtering, among the features kept in each model, immune and epithelial cells were the top-ranked features that predict *ACE2* expression in normal tissues and tumors. Microbiota was observed to be important features for *ACE2* in normal tissues but not in tumors (**Figure 5A** and **5B**). For *TMPRSS2* expression, epithelial cell abundance is the most important predictor in both normal and tumor samples (**Figure 5C** and **5D**). Taken together, these results suggested that immune signatures, epithelial cells, and commensal microbiota were important predictors for *ACE2* expression, while *TMPRSS2* expression was primarily determined by epithelial cells.

**Figure 5.**
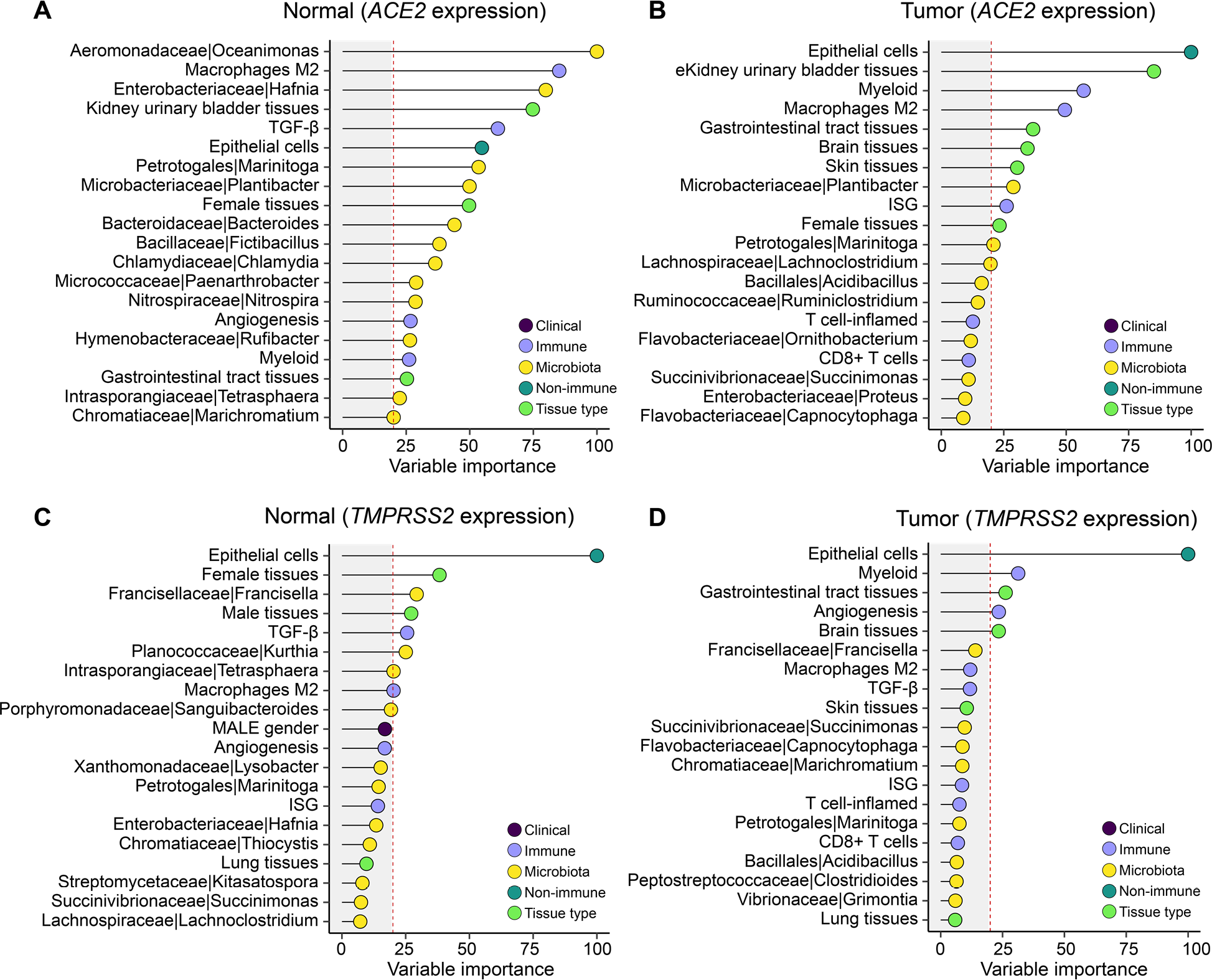
Variable importance of multi-dimensional correlates in predicting *ACE2* and *TMPRSS2* expression. (**A** and **B**) Clinical, immune, and microbiota features in association with *ACE2* gene expression in (**A**) normal and (**B**) tumor samples. (**C** and **D**) Clinical, immune, and microbiota features in association with *TMPRSS2* gene expression in (**C**) normal and (**D**) tumor samples. For each gene, an analysis was performed in normal (*n*=708) and tumor samples (*n*=10,024) separately, with workflow illustrated in **Supplementary Figure 2**. Variable importance scaled to 0-100 is shown on the x-axis. Vertical red dashed line labels score = 20. Features are shown on the y-axis colored by clinical, immune, non-immune, microbiota, and tissue type. Top 20 features ranked by variable importance higher to lower are shown, and the full list is provided in **Supplementary Table 11**. LASSO regression was used in **A** to **D** with 10-fold cross-validation.

## Discussion

We performed a pan-cancer analysis of the receptor that facilitates SARS-CoV-2 infection (ACE2) and the protease that mediates spike protein activation and viral entry (TMPRSS2) by integrating data across six resources including clinical, genetic, transcriptomic and microbiome domains. We found that *ACE2* and *TMPRSS2* are generally expressed lower in tumors relative to matched normal and that digestive organs (both tumor and normal samples) have the highest expression. Neither clinical factors nor HLA genotypes were consistently associated with gene expression levels. Multiple immune gene expression signatures such as ISG and the T cell-inflamed tumor microenvironment did correlate with *ACE2*, and inverse correlations were seen with angiogenesis and TGF-β. *ACE2* expression correlated with increased macrophage abundance in some tumors, while *TMPRSS2* was strongly associated with epithelial cells. Regarding lymphocytes and macrophages, no *ACE2* expression was observed in these cells across multiple single-cell sequencing studies. Microbiota contents are clearly associated with *ACE2* and *TMPRSS2* gene expression levels, possibly suggesting a causal role and the potential to be a modifiable biomarker.

The mortality of COVID-19 disease has been substantially greater than that seen with seasonal influenza and led to the identification of or hypothesis that certain clinical factors may be associated with outcomes. The factors of particular focus included advanced age, BMI, and possibly diabetes or other chronic health conditions such as cardio-pulmonary syndromes and immuno-suppression or cancer [19, 23, 24]. In addition, certain races or ethnicities have experienced greater morbidity and mortality due to pandemic [25]. In our analysis of *ACE2*/*TMPRSS2* gene expression in tumors and matched normal tissues, we observe no consistent association for these factors. Further work would be required to investigate other variables associated with these disease states, such as chronic inflammatory conditions, immuno-suppression, and other disparities that may be contributing factors [39, 40], and cellular context is important to interpret the complexity of those associations [41].

An initial hypothesis when considering the deleterious outcomes for patients with cancer and COVID-19 disease was that cancer tissues themselves might have higher expression of viral entry related genes. We found that gene expression levels did not support this to be the case. Rather cancer tissues broadly have lower expression of *ACE2* and *TMPRSS2*, though the cancers of the digestive tract do have the highest relative level among cancer tissues. This suppressed expression level is consistent with that observed in immuno-oncology gene expression studies [42], in which the T cell-inflamed tumor microenvironment has been observed to be lower in cancer compared with matched normal. *ACE2* has been described as a type I interferon-inducible gene [15]. Across our analysis, we see strong correlations of ACE2 with type I (ISG) and type II (T cell-inflamed) interferon signatures consistent with this.

Observing higher *ACE2* levels in T cell-inflamed tumors does suggest cautious consideration in the administration of cancer immunotherapy during the COVID-19 pandemic, especially in patients with tumors of the aerodigestive tract such as head and neck, lung and colorectal/anal tracts. T cell-inflamed gene expression is strongly correlated with treatment response to checkpoint immunotherapy [43] and has not been associated with immune-related adverse events (irAE) [44]. However, if *ACE2* and *TMPRSS2* levels are high, making viral infection potentially more likely, concomitant treatment with checkpoint blockade may potentially change anti-viral host response [45] or possibly obscure rapidly delineation of symptoms such as fatigue, dyspnea, diarrhea [46] and complicate irAE management, especially given emerging evidence that corticosteroids may worsen COVID-19 disease [47].

Direct infection or dysregulation of immune cell populations is an additional area of concern in patients with cancer and more broadly in infected patients. COVID-19 can manifest with lymphopenia with some autopsy series suggesting lymph node or splenic atrophy [48]. Certainly, dysregulated macrophage activity, with the elaboration of IL-6 and other inflammatory cytokines, is a major component of the disease. Studies have raised the possibility that SARS-CoV-2 infects lymphocytes [17] or macrophages [48], leading to COVID-19 associate findings. In our study, we investigated the expression of *ACE2* and *TMPRSS2* across multiple single-cell sequencing databases encompassing nine independent studies. However, we found no evidence of expression in these cells. It must be noted that the possibility exists that type I interferon may induce *ACE2* expression, which would not be captured in our analysis. We would note, however, that previous studies have not definitively determined that T cells or macrophages are infected by SARS-CoV-2, and direct viral culture from purified cell populations would be needed to confirm this. Additionally, multiple other known pathologies associated with sepsis and extreme illness could explain these lymph node and splenic findings, and few patients with COVID-19 have been documented to have an extreme viremia consistent with what would be required as a pre-requisite to such histologic findings.

Immune responses to cancer and in other settings are increasingly being recognized as influenced by the commensal microbiota [49]. We were, therefore, interested in investigating associations of tissue-based microbiota and ACE2 as a surrogate for the risk of SARS-CoV-2 infection. In our analysis, we found strong correlations of specific bacterial flora and high expression of *ACE2* in COVID-19 related organs, including colorectal and kidney. Particularly in colorectal, where presentations with diarrhea have been widely described, we note a ratio of at least 2:1 of gram-negative bacteria in the bacteria populations significantly associated with elevated *ACE2* expression. A dominance of gram-negative bacteria in the fecal microbiota is assumed at baseline, and yet disequilibrium with an increase of these bacteria is associated with diminished immunological outcomes, especially in immunosuppressed patients [50]. In our study, we have analyzed a heterogeneously collected group of tumors and match normal tissues. However, this observation suggests that further investigation of the commensal microbiome in COVID-19 and possibly that bacterial antibiosis related to coronavirus infection might be of relevance in the future.

We note limitations to our report with the acknowledgment that the use of pre-existing data does not fully capture the complexity of active infection by SARS-CoV-2. Rather we sought to investigate factors correlated with viral cell engagement via ACE2 and viral entry via TMPRSS2 as possible associative risk factors that might be entertained on a clinical or translational level when considering risk for patients with cancer and otherwise of COVID-19. Certainly, there may be virus infection-induced changes that are dynamic. However, we believe our analysis to be the most comprehensive catalog of *ACE2* and *TMPRSS2* correlates to date (34 tumor types from 15 tissue types across 10,038 subjects including both tumor samples and matched normal tissues as well as scRNAseq databases consisting of patients with cancer and healthy donors). We also acknowledge that the microbiota we analyzed were identified from tissue RNAseq data, and the sample collection and preparation of tissue RNAseq was not designed originally to completely rule out potential contamination or confirm the vitality of identified microbes. However, these source data constitute the largest collection of microbiota communities identified from patients with cancer, have previously been used in this manner to build prediction algorithms, and the data were optimized via rigorous methodology to control for noise across the data set [30]. We also note that we are unable in this analysis to comment on respiratory or fecal samples from patients infected with COVID-19 and very much look forward to better understanding the functional mechanisms associated with those commensal and pathogenic microbiota related to COVID-19. Lastly, our work does not determine a causal role of those correlates in driving response or severity of COVID-19 disease and would require further mechanistic studies as well as prospective clinical trials in patients to further develop or investigate interventional approaches.

## Conclusions

We have performed a multi-omic analysis of *ACE2* and *TMPRSS2* gene expression related to clinical, genetic, microbiome covariates associated with COVID-19 infection. We have identified novel commensal microbiome associations and further described interferon associated gene expression patterns in normal and tumor tissues related relevant to SARS-CoV-2 infection. These data will hopefully inform sample collection, future analyses, and treatment of patients with cancer and others infected with COVID-19.

## Data Availability

The data are publicly available and the analyses are placed into GitHub

https://github.com/riyuebao/ACE2_TMPRSS2_multicorrelates

## List of abbreviations

SARS-CoV-2: Severe acute respiratory syndrome coronavirus 2
MERS: Middle East respiratory syndrome
ACE2: Angiotensin-converting enzyme 2
TMPRSS2: Type II transmembrane serine protease
BMI: body mass index
TCGA: The Cancer Genome Atlas
GDC: Genomic Data Commons
RSEM: RNA-Seq by Expectation Maximization
FPKM: Fragments Per Kilobase of transcript per Million mapped reads
WES: whole-exome sequencing
TCGA-CDR: TCGA Pan-Cancer Clinical Data Resource
scRNAseq: single-cell RNAseq
Tukey’s HSD: Tukey’s honest significance test
ISG: interferon-stimulated genes
Tinfl: T cell-inflamed
angio: angiogenesis
TGF-β: transforming growth factor-β

## Declarations

### Availability of data and materials

The original data files were downloaded from Genomic Data Commons (https://portal.gdc.cancer.gov/), Pan-Can Atlas studies (https://gdc.cancer.gov/about-data/publications/pancanatlas), and published studies. Preprocessed data files are provided as supplementary tables. Large-size data objects, as well as additional figures and tables, were deposited to GitHub repository (https://github.com/riyuebao/ACE2_TMPRSS2_multicorrelates). Other data will be provided upon request from the corresponding author.

### Authors’ contributions

RB and JJL conceived the study. JJL supervised the project. RB acquired the data, developed the methodology, performed the computations, and analyzed the data. KH parsed the original clinical XML pages from GDC. KH and LH provided critical feedback on the TCGA data and LASSO regression analysis. RB, KH, LH, and JJL interpreted the results. RB and JJL wrote the manuscript. All authors contributed to the final manuscript.

## Acknowledgments

The authors thank C. Reid for technical assistance on retrieving metadata attributes from GDC.

